# Combination of serum neurofilament light chain and serum cardiac troponin T as biomarkers improves diagnostic accuracy in amyotrophic lateral sclerosis

**DOI:** 10.1101/2025.05.22.25328178

**Authors:** Paula Lindenborn, Rachel Fabian, Torsten Grehl, Huelya Nazlican, Thomas Meyer, Sarah Bernsen, Patrick Weydt

## Abstract

**Objective:** To evaluate the diagnostic performance of serum neurofilament light chain (sNfL) and cardiac troponin T (cTnT) as biomarkers for amyotrophic lateral sclerosis (ALS) and to determine whether their combination improves diagnostic accuracy.

**Methods:** We retrospectively analyzed 293 ALS patients, 47 neurodegenerative disease controls and 24 healthy controls. An independent validation cohort of 501 ALS patients was additionally analyzed to confirm reproducibility of the results. Receiver operating characteristic (ROC) curve analysis was performed for sNfL, cTnT and their combination, and the area under the curve (AUC) was compared across groups. An ALS-specific cTnT cut-off of 8.35 ng/L was determined using the Youden index and applied in subgroup analyses, in which biomarker-negative ALS patients (normal sNfL and cTnT) were compared to biomarker-positive patients regarding disease duration and progression rate.

**Results:** sNfL alone showed excellent performance in discriminating ALS patients from healthy controls (AUC = 0.951), but only moderate performance in discriminating neurodegenerative disease controls (AUC = 0.789). Combining sNfL and cTnT improved diagnostic accuracy for ALS over neurodegenerative disease controls, with a combined AUC of 0.866. Similar AUCs were observed in the validation cohort. Biomarker-negative ALS patients had a longer disease duration (73.0 vs. 18.0 months, p=0.0003) and a lower progression rate (0.19 vs. 0.70 points per months, p<0.0001) than biomarker-positive patients.

**Interpretation:** While sNfL alone performs well in distinguishing ALS from healthy controls, cTnT provides additional value in distinguishing ALS from disease controls. The combination of sNfL and cTnT improves diagnostic accuracy and may help identify clinically distinct ALS subgroups.

## INTRODUCTION

Amyotrophic lateral sclerosis (ALS) is a progressive and ultimately fatal neurodegenerative disease characterized by the selective degeneration of upper motor neurons (UMN) and lower motor neurons (LMN).^1,2^ As the disease progresses, symptoms spread and cause generalized weakness, respiratory impairment, and ultimately death due to respiratory failure.^1^ The complex pathophysiology results in both neuroaxonal damage and neuromuscular changes. ^3^

Neurofilament light chain (NfL) is a structural protein of the neuronal cytoskeleton that is released into the cerebrospinal fluid (CSF) and blood during axonal stress. Elevated levels of NfL in CSF and serum are strongly associated with neurodegeneration and are now recognized as reliable biomarkers for ALS.^4,5,6^. While NfL in CSF has been extensively studied, numerous studies have demonstrated that serum NfL (sNfL) strongly correlates with CSF levels and offers comparable diagnostic performance.^7^. Importantly, sNfL measurement is minimally invasive and more suitable for long-term monitoring, making it a valuable tool for clinical practice. However, sNfL is not specific to ALS, as elevated levels are also observed in other neurodegenerative conditions such as multiple sclerosis, frontotemporal dementia and spinal cord injury .^8,9,10^ This reduces its utility as diagnostic marker, especially when it comes to differentiating ALS from clinically overlapping disorders.

Serum cardiac troponin T (cTnT) is commonly used to detect myocardial injury.^11,12^ However, recent studies have shown that cTnT can also serve as a potential biomarker for skeletal muscle involvement in neurouscular diseases, including ALS.^13,14^

Longitudinal studies have shown that cTnT levels increase as ALS progresses and correlate with functional decline, respiratory impairment, and overall disease severity.^15,16^. Importantly, sNfL and cTnT are not correlated, suggesting that they represent distinct aspects of ALS pathology.^14^.

Given the complementary roles of sNfL and cTnT, we hypothesized that the combination of these biomarkers has the potential to improve diagnostic accuracy in ALS. This dual-marker approach could enhance the differentiation of ALS from related conditions and may help characterize clinical subgroups with specific disease characteristics.

In this study, we evaluated the diagnostic performance of sNfL and cTnT both individually and in combination, by comparing ALS patients with healthy controls and neurodegenerative controls. By combining both biomarkers, we aimed to strengthen diagnostic accuracy and refine the biological understanding of ALS.

## MATERIALS AND METHODS

### Study Design and Population

We included in this retrospective analysis a discovery cohort of 293 ALS patients and two control groups: 47 neurodegenerative disease controls and 24 healthy controls. In addition, an independent validation cohort consisting of 501 ALS patients from the Alfred Krupp Hospital Essen was included to replicate and strengthen the findings of the primary analysis. The ALS discovery cohort included individuals seen between November 2020 and March 2023, while the validation cohort consisted of patients seen between January 2023 and April 2025. All the ALS patients were diagnosed according to established clinical criteria, and key clinical and demographic data were recorded.

The neurodegenerative disease controls group consisted of patients from our outpatient clinic with neurodegenerative disorders other than classic ALS. This group included individuals with primary lateral sclerosis (PLS, n = 8), spinal bulbar muscular atrophy (SBMA, n = 8), Huntington’s disease (HD, n = 28), hereditary sensory and motor neuropathy type 1 (HSMN1, n = 2), and one case of non-choreatic Huntington’s disease.

The healthy controls group included individuals with presymptomatic Huntington’s disease (n = 8), benign fasciculation crampus-syndrome (BFCS, n = 9) and somatoform disorders (n = 7).

For all participants, demographic data (age, sex, Body-Mass-Index (BMI)) as well as serum levels of cTnT and NfL were collected. In ALS patients, functional status was assessed using the ALS Functional Rating Scale (ALSFRS-R), a validated 12-item questionnaire evaluating bulbar, fine motor, gross motor, and respiratory function (score range: 0–48). Disease progression was quantified by calculating the progression rate (PR), defined as: PR = (48 – ALSFRS-R at time of assessment) / disease duration in months. For the validation cohort, the same clinical parameters were collected, including age, ALSFRS-R, disease duration, progression rate and serum NfL and cTnT levels.

As both laboratory measurements and clinical assessments were part of routine diagnostic procedures and analyzed retrospectively, no formal ethical approval was required according to our institutional ethics committee (decision no. 324/20).

### Biomarker Measurements

sNfL was measured using a single-molecule array (Simoa, Quanterix Corporation, Lexington, MA, USA) in the academic laboratory of the University Hospital Ulm, Department of Neurology. sNfL serum concentrations were measured in pg/mL and age-dependent cut-off values were applied based on previous studies^17,18,19^:

- < 51 years: 22 pg/mL
- 51–60 years: 34 pg/mL
- 61–70 years: 45 pg/mL
- 71–80 years: 57 pg/mL
- > 80 years: 78 pg/mL

Serum cTnT was analyzed using a highly sensitive electrochemiluminescence immunoassay (ECLIA) in a commercial laboratory (Labor Volkmann, Karlsruhe, Germany). cTnT serum concentrations were measured in ng/L and to determine the optimal diagnostic cut-off value for cTnT in ALS, the Youden index (J = sensitivity + specificity – 1) was calculated from the ROC analysis.

### Statistical Analysis

Normality of data distributions was assessed using the D’Agostino-Pearson omnibus test. As none of the variables showed consistent normal distribution across all groups, continuous variables (such as age, BMI and biomarker concentrations) are uniformly reported as median and interquartile range (IQR). Group comparisons between ALS patients, neurodegenerative disease controls, and healthy controls were conducted using the Kruskal–Wallis test. Differences in disease duration, progression rate, and other clinical parameters regarding to the analysis of the biomarker-negative and - positive ALS patients were analyzed using the Mann–Whitney U test.

Receiver operating characteristic (ROC) curves were generated to evaluate the diagnostic performance of sNfL, cTnT and their combination. ROC analysis was performed for:

- ALS cohort/validation cohort vs. healthy controls
- ALS cohort/validation cohort vs. neurodegenerative disease controls
- ALS cohort/validation cohort vs. all controls combined

The area under the curve (AUC) was calculated to assess diagnostic accuracy. Statistical significance was defined as p < 0.05. To evaluate the combined performance of both biomarkers, a composite score was generated using logistic regression based on sNfL and cTnT levels. The resulting predicted probabilities were used to generate a ROC curve, which was then compared to the individual ROC curves of sNfL and cTnT.

All statistical analyses were performed with GraphPad Prism 10.2.2 (GraphPad Software Inc., San Diego, USA). Data were stored and managed using Microsoft Excel 2019® (Microsoft Redmond, Washington, USA)

## RESULTS

The clinical and demographic characteristics of ALS patients, neurodegenerative disease controls, and healthy controls are summarized in Table 1. The median age of ALS patients was 67.0 years (IQR: 59.0-74.5), which was significantly higher than that of the neurodegenerative disease controls group (59.0 years, IQR: 55.0-67.0) and the healthy controls group (53.5 years, IQR: 44.3-58.8, p < 0.0001). Similarly, the median BMI in the ALS group was 24.2 kg/m^2^ (IQR: 22.1-26.8), which was lower than in the neurodegenerative disease controls group (25.4 kg/m^2^, IQR: 23.0-27.6) and the healthy controls group (29.6 kg/m^2^, IQR: 24.9-32.3, p = 0.0457).

**Table 1.** Summary of patient characteristics.

|  | ALS cohort | ND controls | Healthy controls | P value | Validation cohort |
| --- | --- | --- | --- | --- | --- |
| **n** | 293 | 47 | 24 | NA | 501 |
| **Sex, female, n (%)** | 127 (43.3%) | 19 (40.4%) | 12 (50%) | NA | 203 (40.5%) |
| **Age, years** | 67.0<br>(59.0-74.5) | 59.0<br>(55.0-67.0) | 53.5<br>(44.3-58.8) | <0.0001 | 63.5<br>(56.0-71.0)<br>n=490 |
| **BMI (kg/m<sup>2</sup>)** | 24.2<br>(22.1-26.8)<br>n=234 | 25.4<br>(23.0 -27.6)<br>n=15 | 29.6<br>(24.9-32.3)<br>n=9 | 0.0457 | NA |
| **ALSFRS-R** | 35.0<br>(29.0-40.0)<br>n=243 | NA | NA | NA | 35.0<br>(28.0-41.0)<br>n=494 |
| **disease duration** | 18.0<br>(9.25-32.8)<br>n=284 | NA | NA | NA | 7.00<br>(2.00-25.5)<br>n=489 |
| **progression rate,**<br>**points per month** | 0.67<br>(0.34-1.22)<br>n=240 | NA | NA | NA | 1.33<br>(0.52-3.33)<br>n=455 |
| **cTnT median**<br>**value (ng/L)** | 18.2<br>(9.00 -33.2) | 6.1<br>(3.90-11.0) | 4.45<br>(3.00-7.58) | <0.0001 | 20.0<br>(11.0-36.0) |
| **sNfL median value**<br>**(pg/mL)** | 93.0<br>(55.5-149.5) | 45.0<br>(25.0-62.0) | 12.5<br>(9.00-29.8) | <0.0001 | 51.5<br>(27.8- 96.1) |

All values are presented as median (Interquartile range). P values refer to comparisons between ALS patients, neurodegenerative disease controls and healthy controls (Kruskal Wallis test). ALS, amyotrophic lateral sclerosis; ALSFRS-R, revised amyotrophic lateral sclerosis functional rating scale, BMI, body mass index; cTnT cardiac troponin T; Neurodegenerative disease controls, ND controls; sNfL, serum neurofilament light chain.

At baseline, ALS patients had a median ALS Functional Rating Scale-Revised (ALSFRS-R) score of 35.0 (IQR: 29.0-40.0) a median disease duration of 18.0 months (IQR: 9.25-32.8) and a median progression rate of 0.67 points per month (IQR: 0.34-1.22). The median sNfL level in ALS patients was 93.0 pg/mL (IQR: 55.5-149.5), compared to 45.0 pg/mL (IQR: 25.0-62.0) in the neurodegenerative disease controls group and 12.5 pg/mL (IQR: 9.00-29.8) in the healthy controls group (p < 0.0001). Similarly, serum cTnT levels were significantly higher in ALS patients (18.2 ng/L, IQR: 9.00-33.2) compared to neurodegenerative disease controls (6.1 ng/L, IQR: 3.90-11.0) and healthy controls (4.45 ng/L, IQR: 3.00-7.58, p < 0.0001). To confirm the robustness of our findings, we also analyzed an independent validation cohort (n_v_ = 501). Compared to the discovery cohort, patients in the validation cohort were slightly younger (median age 63.5 years, IQR: 56.0-71.0 and had a shorter disease duration (7.0 months, IQR: 2.0-25.5). The median progression rate was higher in the validation cohort (1.33 points per month, IQR: 0.52-3.33). In contrast, ALSFRS-R scores at baseline were similar between cohorts (median 35.0 in both groups). Median serum cTnT levels were comparable between the two cohorts (20 ng/L, IQR: 11.0-36.0), whereas median sNfL levels were lower in the validation cohort (51.5 ng/mL, IQR: 27.8-96.1) (*Table 1*).

In the context of myocardial assessment, a pathological threshold of 14 ng/L is conventionally used to discern between elevated and non-elevated serum cTnT levels.^11^ In the present study, a modified cut-off value was determined using the best Youden index derived from ROC analysis. The optimal threshold was 8.35 ng/L (Youden index = 0.511). Using the conventional cardiac cut-off, 92.2 % of ALS patients were identified as biomarker-positive. With the new disease-specific cut-off, this proportion increased to 96.3%. Applying this lower cut-off led to the identification of 12 additional ALS patients (4.1%) with elevated cTnT levels who would have remained biomarker-negative using the conventional cardiac reference, thereby improving diagnostic sensitivity (*Figure 1A, B*). To further validate this threshold, we combined the discovery cohort with the independent validation cohort (n=794) and performed ROC analysis for cTnT as well. The optimal cut-off identified in this combined dataset was again 8.35ng/L, yielding a Youden index of 0.5614, thus supporting the robustness and reproducibility of the proposed ALS-specific threshold. Applying this threshold to the validation cohort, we classified 34 additional ALS patients (6.8%) as biomarker-positive - a slightly higher proportion compared to the discovery cohort *(Figure 1C, D*).

**Figure 1.**
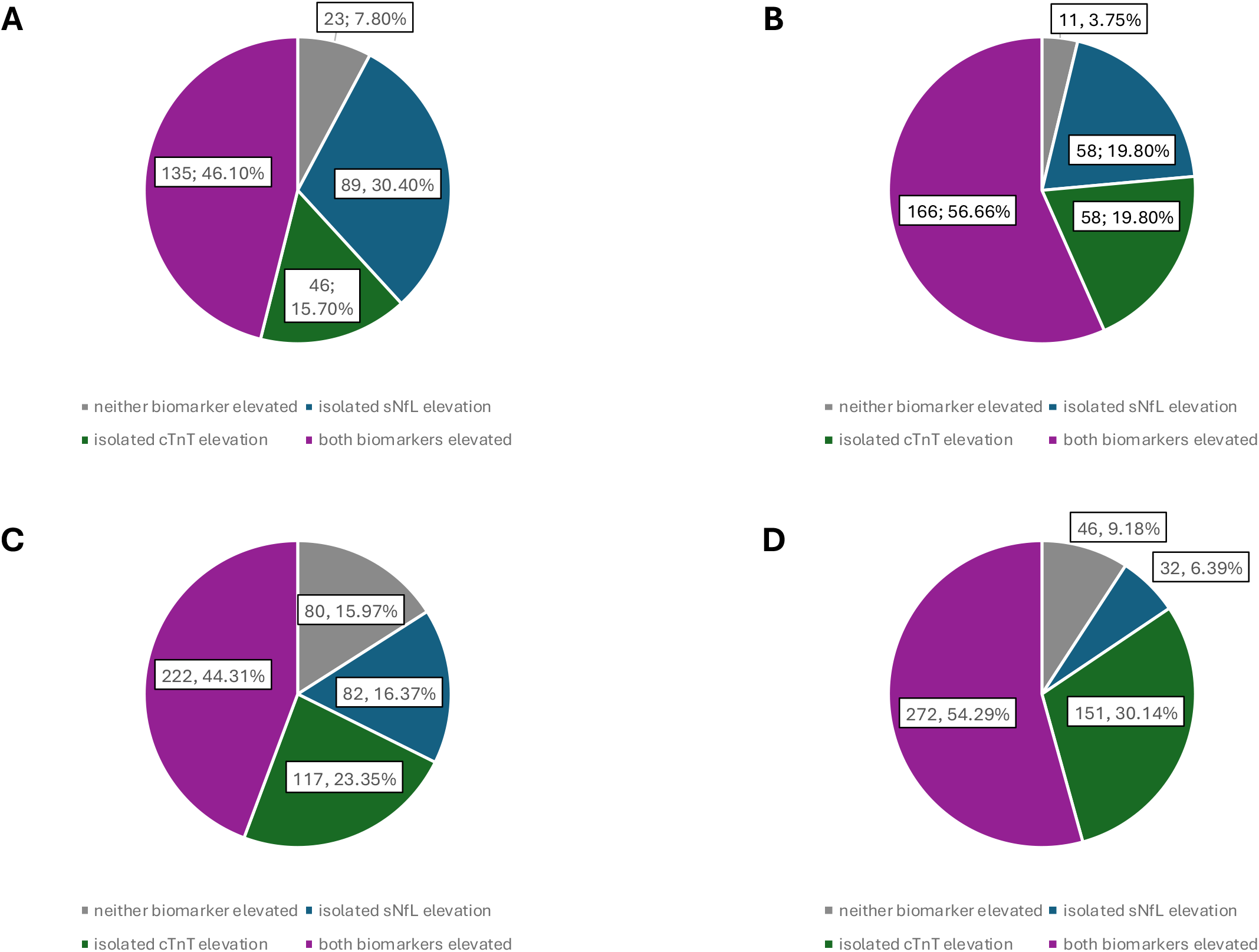
Distribution of biomarker status in ALS patients of the discovery and the validation cohort using two different cTnT cut-offs. (**A**) and (**B**) show classifications in the **discovery cohort** using the conventional cardiac cut-off (14 ng/L) and the ALS-specific cut-off (8.35 ng/L). (**C**) and (**D**) show the corresponding classifications in the **validation cohort** using the same two cut-offs. In all diagrams, ALS patients are stratified into four subgroups based on biomarker status: neither biomarker elevated (normal sNfL and cTnT; isolated sNfL elevation (elevated sNfL, normal cTnT); isolated cTnT elevation (elevated cTnT, normal sNfL) both biomarkers elevated (sNfL and cTnT elevated); Percentages refer to the proportion of patients in each subgroup relative to the entire ALS discovery cohort (n=293) or the validation cohort (n=501). The application of the ALS specific cTnT cut-off (B) and (D) reduced the number of biomarker-negative patients and increased the proportion of patients with either isolated or combined biomarker elevations.

When comparing ALS patients of the discovery cohort with healthy controls, serum NfL achieved an AUC of 0.9508 (95% CI: 0.9210–0.9806, p < 0.0001), while cTnT alone yielded an AUC of 0.8333 (95% CI: 0.7478–0.9187, p < 0.0001). When combined, sNfL and cTnT produced an AUC of 0.9675 (95% CI: 0.9453–0.9898, p < 0.0001), which was higher than the AUCs of the individual biomarkers. When comparing these ALS patients with neurodegenerative disease controls, serum NfL alone reached an AUC of 0.7888 (95% CI: 0.7332–0.8445, p < 0.0001), and cTnT alone yielded an AUC of 0.7612 (95% CI: 0.6869–0.8354, p < 0.0001). The combined use of sNfL and cTnT resulted in an AUC of 0.8657 (95% CI: 0.8221–0.9092, p < 0.0001).). When ALS patients were compared with all control subjects combined, the AUC for the combined biomarkers was 0.8981 (95% CI: 0.8646–0.9317, p < 0.0001) (*Figure. 2*). These results were further supported by data from the validation cohort, which showed a largely consistent diagnostic performance. When comparing ALS patients with healthy controls, sNfL achieved an AUC of 0.8519 in the validation cohort (vs. 0.9508 in the discovery cohort), and cTnT reached and AUC of 0.8892 (vs. 0.8333). The combination of both biomarkers showed an AUC of 0.9105 (vs. 0.9675). In the comparison with neurodegenerative disease controls, sNfL achieved an AUC of 0.5861 (vs. 0.7888), cTnT an AUC of 0.8189 (vs. 0.7612) and the combination 0.8074 (vs. 0.8657). When compared to all controls combined the AUCs in the validation cohort were 0.6760 for sNfL (vs. 08436), 0.8426 for cTnT (vs. 0.7855) and 0.8364 (vs. 0.8981) for combined biomarkers (*Figure 3*).

**Figure 2.**
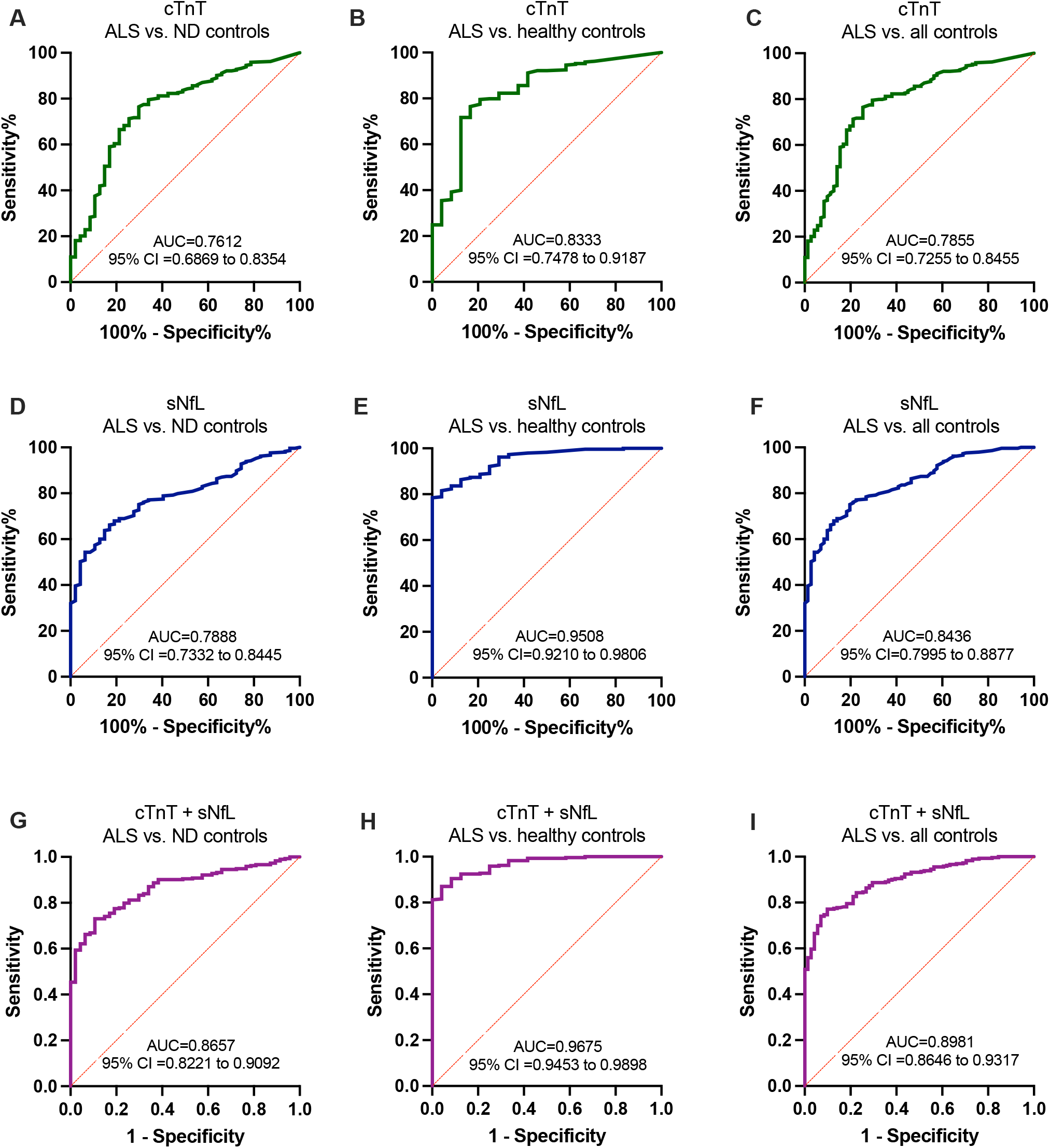
ROC Curves of cTnT, sNfL and thier combination in the discovery cohort with a calculated Area under the Curve (AUC) and 95% confident interval (95% CI) (**A**) ROC curve illustrates cTnT (green line) association with ALS diagnosis in comparison to neurodegenerative disease controls (ND controls) ((**B**) ROC curve illustrates cTnT (green line) association with ALS diagnosis in comparison to healthy controls (**C**) ROC curve illustrates cTnT (green line) association with ALS diagnosis in comparison to all controls (**D**) ROC curve illustrates sNfL (blue line) association with ALS diagnosis in comparison to ND controls (**E**) ROC curve illustrates sNfL (blue line) association with ALS diagnosis in comparison healthy controls (**F**) ROC curve illustrates sNfL (blue line) association with ALS diagnosis in comparison to all controls (**G**) ROC curve illustrates cTnT + sNfL (purple line) association with ALS diagnosis in comparison to ND controls (**H**) ROC curve illustrates cTnT + sNfL (purple line) association with ALS diagnosis in comparison to healthy controls (**I**) ROC curve illustrates cTnT + sNfL (purple line) association with ALS diagnosis in comparison to all controls. All curves were calculated with p value of <0.0001.

**Figure 3.**
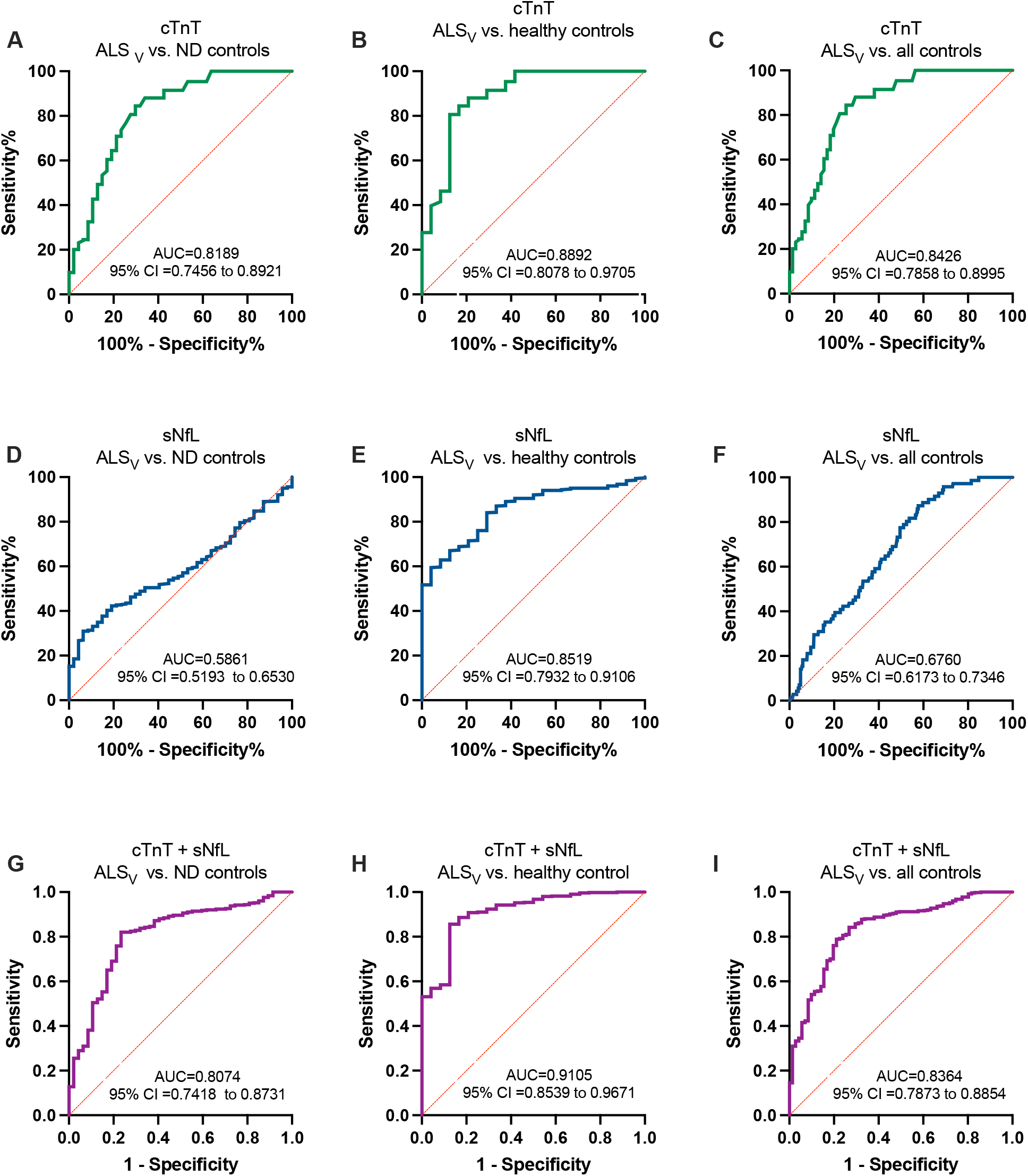
ROC Curves of cTnT, sNfL and their combination in the validation cohort with a calculated Area under the Curve (AUC) and 95% confident interval (95% CI) (**A**) ROC curve illustrates cTnT (green line) association with ALS diagnosis in comparison to neurodegenerative disease controls (ND controls) ((**B**) ROC curve illustrates cTnT (green line) association with ALS diagnosis in comparison to healthy controls (**C**) ROC curve illustrates cTnT (green line) association with ALS diagnosis in comparison to all controls (**D**) ROC curve illustrates sNfL (blue line) association with ALS diagnosis in comparison to ND controls (p value p=0.05) (**E**) ROC curve illustrates sNfL (blue line) association with ALS diagnosis in comparison healthy controls (**F**) ROC curve illustrates sNfL (blue line) association with ALS diagnosis in comparison to all controls (**G**) ROC curve illustrates cTnT + sNfL (purple line) association with ALS diagnosis in comparison to ND controls (**H**) ROC curve illustrates cTnT + sNfL (purple line) association with ALS diagnosis in comparison to healthy controls (**I**) ROC curve illustrates cTnT + sNfL (purple line) association with ALS diagnosis in comparison to all controls. All curves were calculated with p value of <0.0001, unless otherwise is stated.

In addition, we specifically analyzed the subgroup of ALS patients with normal levels of both biomarkers (biomarker-negative; neither sNfL nor cTnT elevated) and compared them to ALS patients with elevated biomarker levels (biomarker-positive; sNfL and/or cTnT elevated). Biomarker-negative ALS patients were rare (n = 11, 3.8%) and exhibited a significantly longer median disease duration (73.0 months, IQR: 26.0-129.0)) than biomarker-positive patients (n = 282, 18.0 months, IQR: 9.00-31.0, *p* = 0.0003). Similarly, the biomarker-negative group had a significant slower median progression rate of 0.19 points per month (IQR: 0.06-0.36), compared to 0.70 points per month (IQR: 0.36-1.25) in biomarker-positive patients (*p* < 0.0001) (*Table 2*). We also conducted a subgroup analysis within the validation cohort comparing biomarker-positive and biomarker-negative ALS patients. Although disease duration did not show a significant difference between these subgroups, biomarker-negative patients (n=46, 9.2%) exhibited a significantly lower progression rate and higher ALSFRS-R score (*Table 2*).

**Table 2.**
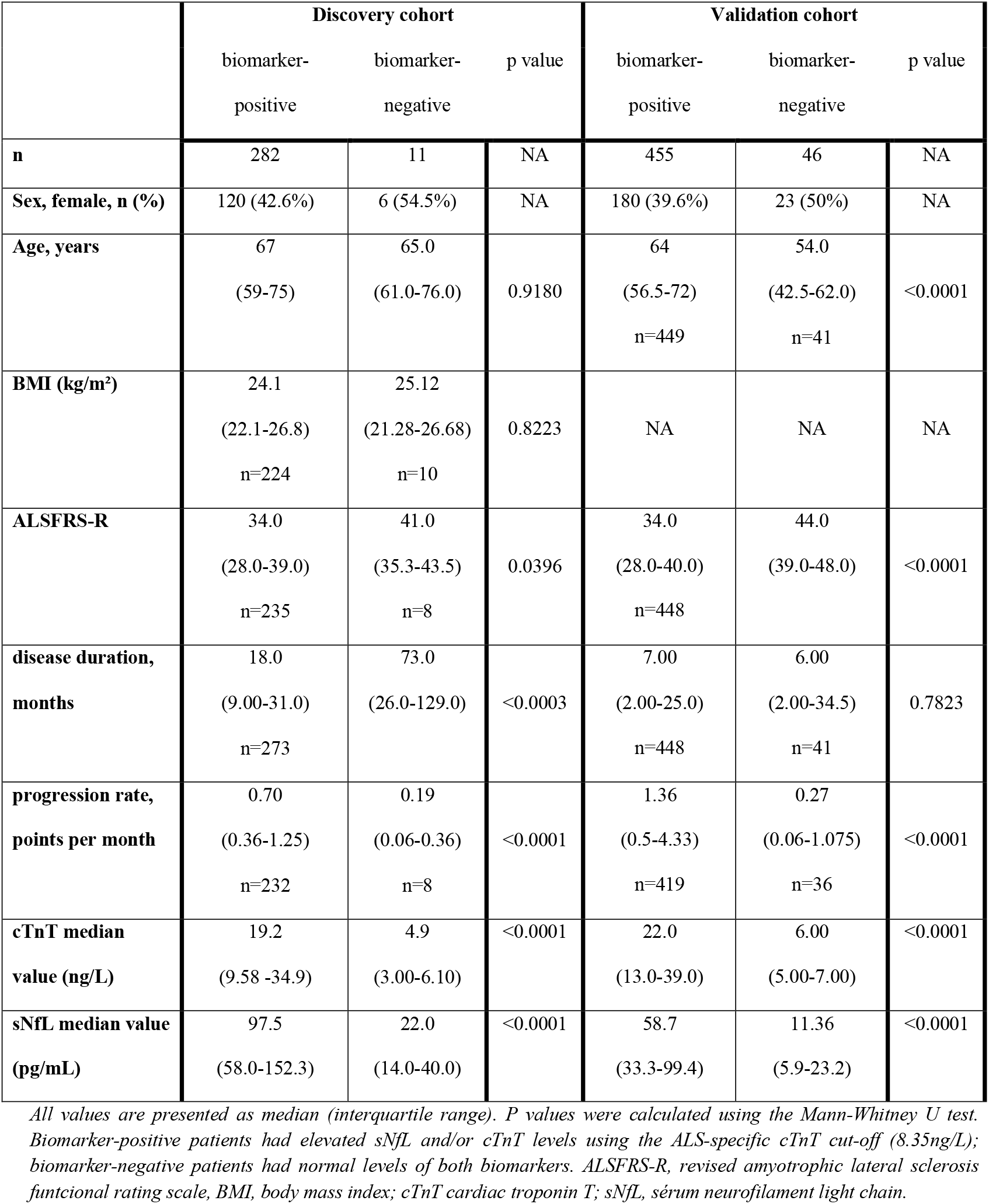
Comparison between biomarker-positive and biomarker-negative ALS patients in the discovery and the validation cohort.

|  | Discovery cohort |  |  | Validation cohort |  |  |
| --- | --- | --- | --- | --- | --- | --- |
|  | biomarker-positive | biomarker-negative | p value | biomarker-positive | biomarker-negative | p value |
| **n** | 282 | 11 | NA | 455 | 46 | NA |
| **Sex, female, n (%)** | 120 (42.6%) | 6 (54.5%) | NA | 180 (39.6%) | 23 (50%) | NA |
| **Age, years** | 67<br>(59-75) | 65.0<br>(61.0-76.0) | 0.9180 | 64<br>(56.5-72)<br>n=449 | 54.0<br>(42.5-62.0)<br>n=41 | <0.0001 |
| **BMI (kg/m<sup>2</sup>)** | 24.1<br>(22.1-26.8)<br>n=224 | 25.12<br>(21.28-26.68)<br>n=10 | 0.8223 | NA | NA | NA |
| **ALSFRS-R** | 34.0<br>(28.0-39.0)<br>n=235 | 41.0<br>(35.3-43.5)<br>n=8 | 0.0396 | 34.0<br>(28.0-40.0)<br>n=448 | 44.0<br>(39.0-48.0) | <0.0001 |
| **disease duration, months** | 18.0<br>(9.00-31.0)<br>n=273 | 73.0<br>(26.0-129.0) | <0.0003 | 7.00<br>(2.00-25.0)<br>n=448 | 6.00<br>(2.00-34.5)<br>n=41 | 0.7823 |
| **progression rate, points per month** | 0.70<br>(0.36-1.25)<br>n=232 | 0.19<br>(0.06-0.36)<br>n=8 | <0.0001 | 1.36<br>(0.5-4.33)<br>n=419 | 0.27<br>(0.06-1.075)<br>n=36 | <0.0001 |
| **cTnT median value (ng/L)** | 19.2<br>(9.58 -34.9) | 4.9<br>(3.00-6.10) | <0.0001 | 22.0<br>(13.0-39.0) | 6.00<br>(5.00-7.00) | <0.0001 |
| **sNfL median value (pg/mL)** | 97.5<br>(58.0-152.3) | 22.0<br>(14.0-40.0) | <0.0001 | 58.7<br>(33.3-99.4) | 11.36<br>(5.9-23.2) | <0.0001 |

*All values are presented as median (interquartile range). P values were calculated using the Mann-Whitney U test. Biomarker-positive patients had elevated sNfL and/or cTnT levels using the ALS-specific cTnT cut-off (8.35ng/L); biomarker-negative patients had normal levels of both biomarkers. ALSFRS-R, revised amyotrophic lateral sclerosis functional rating scale, BMI, body mass index; cTnT cardiac troponin T; sNfL, serum neurofilament light chain.*

## DISCUSSION

In this study, we confirmed that ALS patients exhibit elevated levels of both serum neurofilament light chain (sNfL) and cardiac troponin T (cTnT). Individually, each biomarker demonstrated good diagnostic accuracy, but their combination led to a substantial increase in discriminatory performance, and this improvement was particularly pronounced when distinguishing ALS patients from the neurodegenerative controls group.

Consistent with previous literature, we observed excellent discriminatory power of sNfL for differentiating ALS from healthy controls.^4,19^ However, when comparing ALS with neurodegenerative disease controls, the performance of sNfL alone was reduced, highlighting a known limitation of its diagnostic specificity. This observation aligns with earlier studies showing variable discriminative performance depending on the control cohort. ^5^ Building on this, our validation cohort provided additional support for the observed limitations of sNfL and the complementary value of cTnT. Despite demographic and clinical differences – such as younger age, shorter disease duration and higher progression rate – the main findings were reproducible. While sNfL continued to show good discrimination between ALS patients and healthy controls, its performance in differentiating ALS from neurodegenerative disease controls was notably lower in the validation cohort (AUC = 0.5861) than in the discovery cohort (AUC = 0.7888). This difference further illustrates the limitations of using sNfL alone for differential diagnosis in this context.

The additional integration of cTnT—conventionally a marker of myocardial injury, but increasingly recognized as relevant in skeletal muscle pathology—substantially enhanced diagnostic performance. These findings are in line with results reported by Castro-Gomez et al.^14^, who observed cTnT elevations in ALS, and by Kläppe et al.^15^, although the latter did not report a significant gain from combining biomarkers, potentially due to differences in control group composition and sample size. The comparative study by Vidovic et al.^3^ also highlights the growing importance of muscle damage markers as a complement to neurofilaments in the biomarker profiling of ALS. This further supported by our validation cohort, in which cTnT alone showed consistent performance across control groups and the combination of sNfL and cTnT resulted in improved diagnostic accuracy compared to either marker alone.

Importantly, based on ROC analysis and the Youden index, we identified an optimized diagnostic cut-off for cTnT at 8.35 ng/L - well below the conventional cardiac reference limit of 14.0 ng/L.^11^ This lower threshold may allow for earlier identification of ALS-related skeletal muscle involvement. Preliminary data suggest that using this new cut-off may help detect a subset of ALS patients who would otherwise fall below conventional thresholds. In our cohort, 3.5% of ALS patients were newly identified as biomarker-positive when applying the adjusted cut-off, supporting its potential diagnostic relevance. This effect was even more pronounced in the validation cohort, where 6.8% of patients were newly classified as biomarker-positive. Further validation in independent cohorts is warranted.

A key finding of our study was the identification of a small subgroup of biomarker-negative ALS patients (with neither elevated sNfL nor cTnT). These patients, although rare (3.8% in the discovery cohort and 9.2% in the validation cohort), showed significantly slower disease progression, longer disease duration and a higher ALSFRS-R compared to biomarker-positive individuals. This may correspond to a clinically distinct, more slowly progressive ALS subgroup. Our results are in line with earlier studies linking lower sNfL levels to reduced disease aggressiveness and may also support recent findings associating elevated cTnT with respiratory decline and worse prognosis.^6, 16^

In conclusion, our findings support the use of sNfL and cTnT as complementary biomarkers in ALS. Their combined assessment enhances diagnostic accuracy and may enable earlier detection and characterization of distinct subgroups with prognostic implications. The newly proposed cTnT cut-off may represent a valuable diagnostic tool, although further prospective and longitudinal studies are needed to confirm its clinical utility and prognostic significance. Further research should examine the diagnostic stability of this cut-off across diverse cohorts and explore the clinical features of biomarker-negative subtypes, as well as the temporal dynamics of biomarker levels in relation to disease progression and survival.

## Data Availability

All data produced in the present study are available upon reasonable request to the authors.

## Acknowledgement statement (including conflict of interest and funding sources)

The authors have no conflict of interest to declare.

## Notes

### Competing Interest Statement

The authors have declared no competing interest.

### Funding Statement

The research was supported through unrestricted donations from our patients (to TG and PW), especially Bruno Schmidt (1965 2022) and the Initiative Alle Lieben Schmidt e. V. to PW. TG and PW are also grateful for support from the Boris Canessa Foundation.

### Author Declarations

As both laboratory measurements and clinical assessments were part of routine diagnostic procedures and analyzed retrospectively, no formal ethical approval was required according to our institutional ethics committee (decision no. 324/20 Institutional Ethics Review Board University Hospital Bonn).

